# Trends in prevalence and determinants of unintended pregnancy among Bangladeshi women from 2007 to 2018: A comparative analysis of multiple Demographic Health Surveys

**DOI:** 10.1101/2024.07.10.24310199

**Authors:** Farhana Karim, Abdullah Nurus Salam Khan, Mohiuddin Ahsanul Kabir Chowdhury, Tahmidul Haque, SM Rokonuzzaman, Sharif Uddin Lotus, Sk Masum Billah, Muhammad Sanowar Khan, Md. Shahjahan Siraj

**Author notes:** **Correspondence:** Tahmidul Haque.

## Abstract

**Background:** Unintended pregnancy is a global challenge, particularly prevalent in developing regions, with significant negative impacts on women’s health and well-being. Bangladesh has made progress in fertility decline but still faces challenges due to high rates of unintended pregnancies, unsafe abortions, and limited contraceptive use.

**Methods:** The analysis utilized secondary data from the Bangladesh Demographic and Health Surveys (BDHS) conducted in 2007, 2011, 2014, and 2017-18. The surveys employed a nationally representative sampling frame using a two-stage sampling technique, covering residential households across Bangladesh. The study included 28042 ever-married women aged 15-49 from the four surveys. Descriptive statistics and chi-squared tests examined the relationships between the explanatory and dependent variables. Binary logistic regression was performed to determine the adjusted effects of the selected factors, presenting the results as odds ratios (OR) with 95% confidence intervals (CI). Stata 15 software was used for data analysis, with (p < 0.05) considered statistically significant.

**Results:** The percentage of unintended pregnancies decreased from 29% in 2007 to 21% in 2017-18. Mothers aged 20-29 years had lower likelihood of unintended pregnancy (OR: 0.73-0.81), compared to aged 30 years and above (OR: 1.40). Mothers with secondary education were more likely to have unintended pregnancy in 2007 (OR: 1.28), but less likely in 2011 (OR: 0.75). Employed mothers had higher likelihood of unintended pregnancy (OR: 1.19-1.31), while Muslim mothers had higher likelihood in 2011 and 2014 (OR: 1.33-1.53), but lower likelihood in 2017-18 (OR: 0.73). Unmet need for contraception was consistently associated with higher odds of unintended pregnancy (OR: 2.12-3.94).

**Conclusion:** Unintended pregnancies in Bangladesh have decreased over the past decade, but still pose challenges for women’s reproductive health. Targeted efforts are needed to address factors such as poverty, education, contraception access, and cultural norms to further reduce unintended pregnancies and improve maternal and child well-being.

## Background

Unintended pregnancy refers to pregnancies that are either mistimed (occurred as unplanned), or unwanted (occurred when couples had no desire for more children) [1]. Globally, every year 87 million women experience pregnancy unintentionally [2]. While the unintended pregnancy rate fell worldwide from 1990–1994 to 2010–2014, it dropped less sharply in developing regions (16%) than in developed regions (30%) per 1000 women [3]. The noteworthy slow declining rate of unintended pregnancy also corresponds to a substantial unmet need for contraception in developing regions [4]. A quarter of the unintended pregnancies result in unsafe abortions which is one of the leading causes of maternal mortality [5]. Moreover, poor utilization of maternal health services, poor mental health, low birth weight, preterm births among children and poor breastfeeding practices among mothers are some of the unprecedented consequences of unintended pregnancy [6-9]. These all have negative impacts on women’s quality of life, and at the same time alters mental health status of the women which eventually escalates maternal and neonatal morbidity and mortality [10]. As a significant contributor to maternal and child morbidity and mortality, in resource constrained countries including poor reproductive healthcare infrastructure, unintended pregnancy poses significant barriers in achieving the desired maternal and child health improvements as per Sustainable Development Goal (SDG) indicators.

Bangladesh has shown notable progress in fertility decline, from 6.3 births per woman in the mid-1970s to 2.3 births per woman in 2017-18. However, total wanted fertility rate (TWFR) was 1.7 children in 2017-18, which implies that Bangladeshi women have 0.6 children more than their wanted number of children [11]. Unintended pregnancy often leads to unsafe abortions especially in countries where abortions are illegal and cost women, both financially and in terms of their health and lives [1]. These unsafe abortions result in the death of 80,000 women annually, majority of the burden taking place in developing countries [12, 13]. In recent years unintended pregnancy has been portrayed as a primary indicator of the state of reproductive health in Bangladesh [14, 15]. Majority of the unintended pregnancies are due to non-use or inaccurate use of contraceptives, contraceptive discontinuation, or a noticeable contraceptive failure and lack of family planning awareness [10, 16, 17]. The reported use of contraception is around 80% in developed nations, while the rate still being low in developing nations of Asia (67%) [18]. In the last few decades, Bangladesh has experienced a sharp increase in the contraceptive prevalence, from 7.7% to 62% during the period of 1975 to 2017-18. It is possible that a comparatively lower rate of contraceptive usage might be associated with the increase in unintended pregnancies across the globe, and henceforth in Bangladesh [16, 19]. A variety of other determinants of unintended pregnancy has been cited across the world in different studies. In context of Bangladesh, maternal education, maternal age, wealth index, birth order and age at marriage were found to be associated with unintended pregnancy earlier [20-22]. Particularly, birth order was identified as the single most crucial indicator attributing to unintended pregnancy, followed by mothers’ educational status.

All the previous studies focused on either particular population (i.e., rural communities) or marginalized sections of the society (i.e., mothers from urban slums and other informal settlements) while exploring the factors contributing to unintended pregnancy. Also, these studies used data from separate time points such as Bangladesh Demographic Health Survey (BDHS) 2011 and 2014. But a collective study on a representative population for whole Bangladesh using a series of data from different time points till date has not been conducted. Given that maternal mortality rates in Bangladesh have been stagnant for the last few years (as per Bangladesh Maternal Mortality Survey, 2016), it is high time to investigate all potential causes of maternal mortality, with a particular focus on unwanted pregnancy. In this study an attempt has been offered to investigate the factors which are correlated to unintended pregnancy in Bangladesh using data sets from four BDHS from last 10 years (BDHS 2007, BDHS 2011, BDHS 2014 and BDHS 2017-18). The results of this study will be useful to design and develop programs with impactful strategies for improving reproductive healthcare amenities to reduce unintended pregnancy in Bangladesh.

## Methods

### Data sources

The analysis relies on secondary data sourced from the Bangladesh Demographic and Health Surveys (BDHS) conducted in 2007, 2011, 2014, and the latest one in 2017-18. The samples for 2007 to 2017-18 were nationally representative and the survey used the sampling frame of list of enumeration areas (EAs) from 2001 and 2011 Population and Housing Censuses of the People’s Republic of Bangladesh, provided by Bangladesh Bureau of Statistics. The surveys employed stratified two-stage sampling technique in order to ensure national representativeness [23]. The first stage involved creating a sampling frame that included a list of the primary sampling units (PSUs) or EAs that encompassed the entire nation. Each PSU or EA is further divided into standard size segments, with each segment containing between 100 and 500 households. During this phase, a sample consisting of preset segments is chosen randomly, with the probability of choosing to be proportional to the size of the Enumeration Area (EA), which is determined by the number of households within the EA. The second stage involves data collectors selecting households systematically from a list of previously enumerated households in each EA section and conducting in-person interviews with target populations: women aged 15–49 and males aged 15–64. Nomadic and institutional groups like prisoners and hotel guests are frequently excluded.

The BDHSs covered 10500, 17141, 17300 and 19547 residential households from 2007, 2011, 2014 and 2017-18, respectively. From these sample households 11440, 17749, 17863 and 20127 women who had been previously married were interviewed, respectively. This study included ever-married women aged 15–49 from four surveys: 6754 in 2007, 9851 in 2011, 5963 in 2014, and 6474 in 2017-18 [11, 24-26].

### Dependent variable

The study’s dependent variable is “pregnancy intentions” based on whether women wanted their current pregnancy. The responses are “then”, “later”, and “not at all”. Using the definition of unintended pregnancy as “pregnancies that are either wanted earlier or later than occurred (mistimed) or not needed (unwanted)” (27), we coded these three replies: then=0 ‘intended’; ‘later and not at all’=1 ‘unintended’. All women (15–49) who answered this question are included.

### Explanatory variables

This study explores 14 explanatory variables, divided under three broad characteristics: socio-demographic variables, obstetric history and contraception/family planning(?). Socio-demographic variables are respondents’ current age (less than 20 years, 20-29 years and 30 years or more), education (no formal education, primary complete or below, secondary incomplete, and secondary complete or higher), employment (yes and no), religion (Islam and Others), wealth index (poorest, poorer, middle, richer and richest), place of residence (urban, rural), family size (≤4, 5-6, ≥7), respondents’ age at first birth (<18 years, ≥18 years), respondents’ husbands’ education (no formal education, primary or below, secondary or higher, and Do not know), sex of household head (male, female) and division (Barisal, Chittagong, Dhaka, Khulna, Rajshahi, Rangpur, Sylhet, Mymensingh). Obstetric history and contraception practice included birth order (1, 1+), ever had a terminated pregnancy (yes, no) and unmet need for contraception (currently married women who are not pregnant, not postpartum amenorrhoeic, are regarded fecund and want to postpone their next birth for 2 or more years or stop childbearing altogether but are not using a contraceptive method, or have a mistimed or unwanted current pregnancy (yes, no).

### Statistical analysis

The respondents’ background traits were described using frequency distribution. All explanatory variables and the dependent variable were compared using two-way contingency tables and Pearson’s chi-squared test. We present a statistical model suitable for binary response, namely the binary logistic regression model in multivariate setting, to obtain the adjusted effects of chosen parameters [27]. The odds ratio (OR) and 95% confidence interval (CI) were used to represent the result. Stata 15 was used for data analysis. P-value less than 0.05 was considered as statistically significant in this study.

### Ethical approval

All the analyses were done using publicly available data from demographic health surveys. All the BDHSs were conducted under the authority of the National Institute of Population Research and Training (NIPORT) of the Ministry of Health and Family Welfare, Bangladesh. All participants gave informed consent before taking part in the survey. The data was made publicly available without any personal identifiers. In this study, further ethical approval was not necessary as the analyses were based on the secondary data available in the public domain in anonymised form.

## Results

The percentage of mother in each category of the selected variables for each survey point are presented in Table 1. More than half mothers were of between 20-29 years of their age (60% in 2007, 63% in 2011, 59% in 2014 and 60% in 2017-18). With respect to the highest education level, percentage of mothers having secondary level education or higher has been increasing since 2007 (12%, 12%, 17% and 23% in 2007, 2011, 2014 and 2017-18, respectively). Similarly, percentages of respondents’ husbands’ having secondary or higher level of education have been increased from 36% in 2007 to 52% in 2017-18. More than one third mothers (39%) were employed in 2017-18, which has been slowly increased since 2007 (29%). Nine out of ten mothers were Muslim and in 2007, eight out of ten mothers lived in rural areas, while it decreased over time in 2017-18 (seven out of ten mothers). Across the four BDHSs, the proportion of male as household head showed slight decrease from 91% in 2007 to 87% in 2017-18. Regarding age at first birth of the respondents, the percentage of having child below 18 years has declined from 51% in 2007 to 39% in 2017-18 BDHS. The percentages of mothers having family size less than four has been slightly increased in 2017-18 BDHS (33%) than 2007 (31%). About half of the mothers in all surveys were residing in Dhaka and Chittagong division. Percentage of mothers who were first time pregnant during surveys, has been increased gradually from 30% in 2007 BDHS to 41% in 2017-18. Less than 20% mothers had history of termination of pregnancy in all surveys. Percentage of unmet need for contraception was gradually declined from 2007 to 2017-18 BDHS (23%, 16%, 16% and 14% in 2007, 2011, 2014 and 2017-18, respectively). Table 1 also shows percent distribution of unwanted births among ever married mothers whose most recent pregnancy occurred in five years preceding the survey or who have been currently pregnant. Findings suggest that, the percentage of unintended (mistimed and unwanted together) pregnancies has been gradually decreased since 2007 and it was 29% in 2007 BDHS, 28% in 2011 BDHS, 26% in 2014 BDHS and finally 21% in 2017-18 BDHS.

**Table 1:** Percentage distribution of characteristics of the respondents, 2007, 2011, 2014 and 2017-18 BDHS.

| Characteristics | BDHS 2007,<br>N= 6754 | BDHS 2011,<br>N=9851 | BDHS 2014,<br>N=5963 | BDHS 2017-18,<br>N=6474 |
| --- | --- | --- | --- | --- |

|  | **No. of**<br>**women** | **%** | **No. of**<br>**women** | **%** | **No. of**<br>**women** | **%** | **No. of**<br>**women** | **%** |
| --- | --- | --- | --- | --- | --- | --- | --- | --- |
| **Maternal Age** |  |  |  |  |  |  |  |  |
| Less than 20 years | 1,175 | 17.4 | 1,515 | 15.4 | 1,345 | 22.5 | 1294 | 20.0 |
| 20 to 29 years | 4,048 | 59.9 | 6,236 | 63.3 | 3,507 | 58.8 | 3884 | 60.0 |
| 30 years and above | 1,530 | 22.7 | 2,100 | 21.3 | 1,112 | 18.6 | 1296 | 20.0 |
| **Education** |  |  |  |  |  |  |  |  |
| No formal education | 1,800 | 26.7 | 1,945 | 19.7 | 837 | 14.0 | 400 | 6.2 |
| Primary complete or<br>below | 2,107 | 31.2 | 3,027 | 30.7 | 1680 | 28.2 | 1774 | 27.4 |
| Secondary incomplete | 1,996 | 29.5 | 3,679 | 37.3 | 2445 | 41.0 | 2829 | 43.7 |
| Secondary complete or<br>higher | 834 | 12.4 | 1,199 | 12.2 | 1002 | 16.8 | 1470 | 22.7 |
| Missing | 17 | 0.3 |  |  |  |  |  |  |
| **Employment** |  |  |  |  |  |  |  |  |
| Yes | 1,928 | 28.6 | 1,067 | 10.8 | 1,445 | 24.2 | 2,501 | 38.6 |
| No | 4,818 | 71.3 | 8,783 | 89.2 | 4,509 | 75.6 | 3,970 | 61.3 |
| Missing | 7 | 0.1 | N/A | N/A | 9 | 0.2 | 2 | 0.0 |
| **Husband's education** |  |  |  |  |  |  |  |  |
| No formal education | 2,357 | 34.9 | 2,876 | 29.2 | 1,423 | 23.9 | 866 | 13.4 |
| Primary or below | 1,963 | 29.1 | 2,883 | 29.3 | 1,810 | 30.4 | 2177 | 33.6 |
| Secondary or higher | 2,423 | 35.9 | 4,084 | 41.5 | 2,728 | 45.7 | 3348 | 51.7 |
| Do not know |  |  |  |  |  |  | 17 | 0.3 |
| Missing | 10 | 0.2 | 8 | 0.1 | 3 | 0.1 | 66 | 1.0 |
| **Religion** |  |  |  |  |  |  |  |  |
| Islam | 6,191 | 91.7 | 9,036 | 91.7 | 5,459 | 91.5 | 5,963 | 92.1 |
| Hindu & other | 562 | 8.3 | 815 | 8.3 | 504 | 8.5 | 510 | 7.9 |
| **Wealth index** |  |  |  |  |  |  |  |  |
| Poorest | 1,507 | 22.3 | 2,283 | 23.2 | 1,280 | 21.5 | 1,326 | 20.5 |
| Poorer | 1,466 | 21.7 | 2,040 | 20.7 | 1,102 | 18.5 | 1,344 | 20.8 |
| Middle | 1,331 | 19.7 | 1,929 | 19.6 | 1,097 | 18.4 | 1,248 | 19.3 |
| Richer | 1,264 | 18.7 | 1,874 | 19.0 | 1,150 | 19.3 | 1,306 | 20.2 |
| Richest | 1,184 | 17.5 | 1,724 | 17.5 | 1,115 | 18.7 | 1,250 | 19.3 |
| Missing |  |  |  |  | 219 | 3.7 |  |  |
| **Sex of household head** |  |  |  |  |  |  |  |  |
| Male | 6,141 | 90.9 | 9,108 | 92.5 | 5,472 | 91.8 | 5,633 | 87.0 |
| Female | 613 | 9.1 | 743 | 7.5 | 491 | 8.2 | 840 | 13.0 |
| **Family size** |  |  |  |  |  |  |  |  |
| 4 or less | 2,108 | 31.2 | 3,182 | 32.3 | 2,016 | 33.8 | 2,102 | 32.5 |
| 5 to 6 | 2,295 | 34.0 | 3,463 | 35.2 | 2,021 | 33.9 | 2,272 | 35.1 |
| 7 and above | 2,351 | 34.8 | 3,206 | 32.5 | 1,926 | 32.3 | 2,099 | 32.4 |
| **Age at first birth** |  |  |  |  |  |  |  |  |
| Less than 18 years | 3,470 | 51.4 | 4,785 | 48.6 | 2,449 | 41.1 | 2,539 | 39.2 |
| 18 years and above | 3,284 | 48.6 | 5,066 | 51.4 | 3,515 | 58.9 | 3,935 | 60.8 |
| **Place of residence** |  |  |  |  |  |  |  |  |
| Urban | 1,393 | 20.6 | 2,189 | 22.2 | 1,576 | 26.4 | 1,739 | 26.9 |
| Rural | 5,361 | 79.4 | 7,662 | 77.8 | 4,426 | 74.2 | 4,734 | 73.1 |
| **Division** |  |  |  |  |  |  |  |  |
| Barisal | 428 | 6.3 | 553 | 5.6 | 345 | 5.8 | 366 | 5.7 |
| Chittagong | 1,480 | 21.9 | 2,227 | 22.6 | 1294 | 21.7 | 1362 | 21.0 |
| Dhaka | 2,101 | 31.1 | 3,078 | 31.2 | 2091 | 35.1 | 1663 | 25.7 |
| Khulna | 656 | 9.7 | 888 | 9.0 | 468 | 7.9 | 594 | 9.2 |
| Rajshahi | 1,478 | 21.9 | 1,291 | 13.1 | 589 | 9.9 | 758 | 11.7 |
| Rangpur |  |  | 1,040 | 10.6 | 567 | 9.5 | 667 | 10.3 |
| Sylhet | 611 | 9.1 | 774 | 7.9 | 609 | 10.2 | 511 | 7.9 |
| Mymensingh |  |  |  |  |  |  | 553 | 8.5 |
| **Birth order** |  |  |  |  |  |  |  |  |
| 1 | 2,050 | 30.4 | 3,109 | 31.6 | 1,977 | 40.8 | 2,081 | 40.8 |
| 1+ | 4,703 | 69.6 | 6,742 | 68.4 | 3,987 | 59.2 | 4,393 | 59.2 |
| **Ever had a terminated pregnancy** |  |  |  |  |  |  |  |  |
| Yes | 1,297 | 19.2 | 1,788 | 18.2 | 873 | 14.6 | 1,063 | 16.4 |
| No | 5,443 | 80.6 | 8,063 | 81.8 | 5,090 | 85.4 | 5,411 | 83.6 |
| **Unmet need for contraception** |  |  |  |  |  |  |  |  |
| Yes | 1,545 | 23.1 | 1,541 | 15.6 | 927 | 15.5 | 812 | 13.6 |
| No | 5,208 | 76.9 | 8,170 | 82.9 | 4,978 | 83.5 | 5,596 | 93.8 |
| Missing | 1 | 0.0 | 140 | 1.4 | 58 | 1.0 | 66 | 1.1 |
| **Pregnancy intention status** |  |  |  |  |  |  |  |  |
| Wanted | 4,799 | 71.1 | 7,126 | 72.3 | 4408 | 73.9 | 5,104 | 78.8 |
| Mistimed | 1,004 | 14.9 | 1,474 | 15.0 | 912 | 15.3 | 861 | 13.3 |
| Unwanted | 951 | 14.0 | 1,250 | 12.7 | 643 | 10.8 | 508 | 7.9 |

The findings regarding distribution of percentage of unintended pregnancy among different variable categories are displayed in Table 2. Univariate analysis showed that the factors significantly associated with unintended pregnancy were mother’s education, employment, husband’s education, religion, family size, age at first birth, birth order and unmet need for contraception. Maternal age was significantly associated with unintended pregnancy in 2007, 2011 and 2014, while wealth index was significantly associated in 2011, 2014 and 2017-18. Sex of household head was significantly associated with unintended pregnancy only in 2017-18 and place of residence had a significant association only in 2014. Division was found significant in BDHS 2007, 2011 and 2017-18, while history of termination of pregnancy was found significant in 2007 and 2011 BDHS.

**Table 2.** Percentage distribution of unintended pregnancy among ever-married women age 15–49, by selected background characteristics, 2007, 2011, 2014 and 2017-18 BDHS.

|  | Unintended Pregnancy Status |  |  |  |
| --- | --- | --- | --- | --- |
| Characteristics | BDHS<br>2007, N=<br>6754 | BDHS<br>2011,<br>N=9851 | BDHS<br>2014,<br>N=5963 | BDHS<br>2017-18,<br>N=6474 |
|  | Yes (%) | Yes (%) | Yes (%) | Yes (%) |
| Maternal Age |  |  |  |  |
| Less than 20 years | 24.1 | 23.8 | 21.2 | 21.6 |
| 20 to 29 years | 25.8 | 24.4 | 23.5 | 23.1 |
| 30 years and above | 41.0 | 40.0 | 40.2 | 20.9 |
|  | p<0.001 | p<0.001 | p<0.001 | p= 0.232 |
| **Mothers' Education** |  |  |  |  |
| No formal education | 32.8 | 34.4 | 38.6 | 29.8 |
| Primary complete or below | 30.6 | 31.2 | 31.0 | 25.8 |
| Secondary incomplete | 25.9 | 24.2 | 21.6 | 19.4 |
| Secondary complete or higher | 24.1 | 18.5 | 18.2 | 16.5 |
|  | p<0.001 | p<0.001 | p<0.001 | p<0.001 |
| **Employment** |  |  |  |  |
| Yes | 32.2 | 33.1 | 30.2 | 24.2 |
| No | 27.7 | 23.9 | 24.7 | 19.3 |
|  | p= 0.019 | p= 0.004 | p<0.001 | p<0.001 |
| **Husband's education** |  |  |  |  |
| No formal education | 30.4 | 32.1 | 35.0 | 28.5 |
| Primary or below | 31.0 | 28.6 | 27.5 | 23.0 |
| Secondary or higher | 25.8 | 23.8 | 20.5 | 18.2 |
| Do not know | N/A | N/A | N/A | 11.2 |
|  | p= 0.001 | p<0.001 | p<0.001 | p<0.001 |
| **Religion** |  |  |  |  |
| Islam | 29.7 | 28.4 | 26.8 | 21.7 |
| Hindu & other | 20.7 | 20.0 | 18.0 | 15.1 |
|  | p<0.001 | p<0.001 | p= 0.002 | p= 0.002 |
| **Wealth index** |  |  |  |  |
| Poorest | 30.6 | 32.5 | 36.8 | 23.6 |
| Poorer | 30.4 | 29.1 | 28.5 | 25.5 |
| Middle | 29.9 | 27.3 | 25.1 | 18.9 |
| Richer | 26.4 | 26.5 | 22.2 | 20.9 |
| Richest | 26.6 | 21.2 | 18.5 | 16.4 |
|  | p= 0.182 | p<0.001 | p<0.001 | p<0.001 |
| **Gender of household head** |  |  |  |  |
| Man | 28.6 | 28.0 | 26.3 | 21.6 |
| Woman | 31.9 | 23.9 | 24.2 | 18.2 |
|  | p= 0.461 | p=0.231 | p= 0.372 | p= 0.017 |
| **Family size** |  |  |  |  |
| 4 or less | 20.7 | 20.4 | 21.4 | 17.5 |
| 5 to 6 | 32.1 | 31.9 | 31.5 | 23.9 |
| 7 and above | 33.2 | 30.3 | 25.3 | 21.8 |
|  | p<0.001 | p<0.001 | p<0.001 | p<0.001 |
| **Age at first birth** |  |  |  |  |
| Less than 18 years | 32.3 | 33.1 | 31.8 | 25.9 |
| 18 years and above | 25.4 | 22.5 | 22.1 | 18.1 |
|  | p<0.001 | p<0.001 | p<0.001 | p<0.001 |
| **Place of residence** |  |  |  |  |
| Urban | 31.7 | 26.5 | 20.7 | 22.2 |
| Rural | 28.2 | 28.0 | 27.8 | 20.8 |
|  | p= 0.066 | p= 0.678 | p= 0.002 | p=0.535 |
| **Division** |  |  |  |  |
| Barisal | 33.5 | 28.2 | 24.6 | 23.8 |
| Chittagong | 29.6 | 26.5 | 22.9 | 16.1 |
| Dhaka | 29.8 | 28.1 | 27.0 | 21.1 |
| Khulna | 26.4 | 27.7 | 27.9 | 24.3 |
| Rajshahi | 27.0 | 30.3 | 29.6 | 19.6 |
| Rangpur | N/A | 27.6 | 24.3 | 21.4 |
| Sylhet | 28.4 | 24.3 | 27.6 | 26.8 |
| Mymensingh | N/A | N/A | N/A | 23.1 |
|  | p= 0.179 | p=0.037 | p=0.017 | p<0.001 |
| **Birth order** |  |  |  |  |
| 1 | 14.3 | 12.5 | 13.4 | 9.6 |
| 1+ | 35.3 | 34.7 | 32.4 | 26.6 |
|  | p<0.001 | p<0.001 | p<0.001 | p<0.001 |
| **Ever had a terminated pregnancy** |  |  |  |  |
| Yes | 30.8 | 30.0 | 26.5 | 22.4 |
| No | 28.4 | 27.2 | 26.0 | 20.9 |
|  | p= 0.007 | p= 0.012 | p= 0.517 | p= 0.193 |
| **Unmet need for contraception** |  |  |  |  |
| Yes | 50.9 | 39.7 | 45.6 | 28.9 |
| No | 22.4 | 25.4 | 22.3 | 20.1 |
|  | p<0.001 | p<0.001 | p<0.001 | p<0.001 |

Table 3 presents the findings of adjusted logistic regressions which revealed that, mothers aged 20 to 29 years were less likely to have unintended pregnancy than mothers below 20 years in 2007 (OR: 0.73, 95% CI: 0.61 - 0.88), 2011(OR: 0.70, 95% CI: 0.60 – 0.82) and 2014 (OR: 0.81, 95% CI: 0.67 - 0.97); however, in 2017-18 it was found that, mothers aged 30 years and above were more likely to have unintended pregnancy than mothers below 20 years (OR: 1.40, 95% CI: 1.11- 1.76).

**Table 3:** Odds ratios (OR) with 95% confidence intervals (CI) of unintended pregnancy and selected explanatory variables among ever-married women in Bangladesh, from 2007, 2011, 2014 and 2017-18 BDHSs.

| Characteristics | BDHS 2007 |  | BDHS 2011 |  | BDHS 2014 |  | BDHS 2017-18 |  |
| --- | --- | --- | --- | --- | --- | --- | --- | --- |
|  | OR | 95% CI | OR | 95% CI | OR | 95% CI | OR | 95% CI |
| **Maternal Age** |  |  |  |  |  |  |  |  |
| Less than 20 years | 1 |  | 1 |  | 1 |  | 1 |  |
| 20 to 29 years | 0.73** | 0.61 - 0.88 | 0.70*** | 0.60-0.82 | 0.81* | 0.67-0.97 | 0.90 | 0.74-1.10 |
| 30 years and above | 1.11 | 0.89-1.38 | 1.09 | 0.90-1.31 | 1.19 | 0.94-1.50 | 1.40** | 1.11-1.76 |
| **Education** |  |  |  |  |  |  |  |  |
| No formal education | 1 |  | 1 |  | 1 |  | 1 |  |
| Primary complete or below | 1.15 | 0.98-1.35 | 1.09 | 0.94-1.26 | 1.05 | 0.85-1.30 | 0.98 | 0.75-1.23 |
| Secondary incomplete | 1.15 | 0.95-1.40 | 1.00 | 0.85-1.19 | 0.81 | 0.64-1.02 | 0.94 | 0.71-1.24 |
| Secondary complete or higher | 1.28* | 1.01-1.63 | 0.75* | 0.60-0.95 | 0.79 | 0.59-1.06 | 1.04 | 0.75-1.44 |
| **Employment** |  |  |  |  |  |  |  |  |
| No | 1 |  | 1 |  | 1 |  | 1 |  |
| Yes | 1.19* | 1.04-1.37 | 1.31** | 1.12-1.53 | 1.19* | 1.02-1.39 | 1.06 | 0.92-1.22 |
| **Husband's**<br>**education** |  |  |  |  |  |  |  |  |
| No formal<br>education | 1 |  | 1 |  | 1 |  | 1 |  |
| Primary or<br>below | 1.09 | 0.93-1.26 | 1.03 | 0.90-1.18 | 1.00 | 0.83-1.20 | 0.91 | 0.74-1.11 |
| Secondary or<br>higher | 1.00 | 0.83-1.19 | 1.09 | 0.94-1.27 | 0.90 | 0.74-1.11 | 0.87 | 0.70-1.08 |
| Do not know |  |  |  |  |  |  | 0.43 | 0.09-1.98 |
| **Religion** |  |  |  |  |  |  |  |  |
| Islam | 1.23 | 0.97-1.56 | 1.53*** | 1.26-1.86 | 1.33* | 1.02-1.73 | 0.73* | 0.56-0.95 |
| Hindu & other | 1 |  | 1 |  | 1 |  | 1 |  |
| **Wealth index** |  |  |  |  |  |  |  |  |
| Poorest | 1 |  |  |  | 1 |  | 1 |  |
| Poorer | 1.03 | 0.85-1.24 | 0.89 | 0.76-1.03 | 0.87 | 0.71-1.06 | 1.19 | 0.98-1.44 |
| Middle | 1.12 | 0.92-1.37 | 0.81* | 0.67-0.96 | 0.90 | 0.73-1.12 | 0.90 | 0.73-1.12 |
| Richer | 1.03 | 0.83-1.27 | 0.82* | 0.69-0.97 | 0.86 | 0.69-1.08 | 0.97 | 0.77-1.21 |
| Richest | 0.88 | 0.68-1.13 | 0.64*** | 0.52-0.79 | 0.88 | 0.67-1.15 | 0.62** | 0.47-0.82 |
| **Gender of household head** |  |  |  |  |  |  |  |  |
| Man | 1 |  | 1 |  | 1 |  | 1 |  |
| Woman | 0.86 | 0.69-1.05 | 0.83 | 0.68-1.02 | 0.74* | 0.58-0.94 | 0.70** | 0.57-0.87 |
| **Family size** |  |  |  |  |  |  |  |  |
| 4 or less | 1 |  | 1 |  | 1 |  | 1 |  |
| 5 to 6 | 1.45 | 1.24-1.69 | 1.54*** | 1.36-1.74 | 1.47*** | 1.25-1.73 | 1.33*** | 1.14-1.57 |
| 7 and above | 1.59 | 1.36-1.85 | 1.88*** | 1.62-2.12 | 1.45*** | 1.22-1.72 | 1.45*** | 1.22-1.72 |
| **Age at first birth** |  |  |  |  |  |  |  |  |
| Less than 18 years | 1 |  | 1 |  | 1 |  | 1 |  |
| 18 years and above | 0.72**<br>* | 0.63-0.81 | 0.70*** | 0.63-0.78 | 0.69*** | 0.60-0.79 | 0.73*** | 0.64-0.85 |
| **Place of residence** |  |  |  |  |  |  |  |  |
| Urban | 1 |  | 1 |  | 1 |  | 1 |  |
| Rural | 0.69**<br>* | 0.58-0.81 | 0.77*** | 0.67-0.89 | 0.94 | 0.80-1.10 | 0.72*** | 0.62-0.85 |
| **Division** |  |  |  |  |  |  |  |  |
| Barisal | 1 |  | 1 |  | 1 |  | 1 |  |
| Chittagong | 0.86 | 0.66-1.11 | 0.75* | 0.60-0.93 | 0.82 | 0.64-1.05 | 0.62** | 0.47-0.82 |
| Dhaka | 0.97 | 0.75-1.25 | 0.93 | 0.75-1.16 | 0.92 | 0.71-1.20 | 0.98 | 0.74-1.29 |
| Khulna | 0.97 | 0.72-1.29 | 1.08 | 0.86-1.37 | 1.33* | 1.01-1.74 | 1.15 | 0.89-1.54 |
| Rajshahi | 0.93 | 0.71-1.22 | 1.01 | 0.80-1.28 | 1.37* | 1.04-1.79 | 0.95 | 0.71-1.27 |
| Rangpur |  |  | 0.96 | 0.76-1.21 | 0.88 | 0.67-1.16 | 1.22 | 0.92-1.61 |
| Sylhet | 0.68** | 0.52-0.90 | 0.66*** | 0.52-0.83 | 0.80 | 0.62-1.04 | 0.92 | 0.69-1.21 |
| Mymensingh |  |  |  |  |  |  | 0.77 | 0.58-1.02 |
| **Birth order** |  |  |  |  |  |  |  |  |
| 1 | 1 |  | 1 |  | 1 |  | 1 |  |
| 1+ | 2.97**<br>* | 2.52-3.50 | 3.58*** | 3.11-4.12 | 2.91*** | 2.43-3.49 | 3.49*** | 2.89-4.22 |
| **Ever had a terminated pregnancy** |  |  |  |  |  |  |  |  |
| No | 1 |  | 1 |  | 1 |  | 1 |  |
| Yes | 1.06 | 0.92-1.23 | 0.99 | 0.87-1.12 | 0.97 | 0.81-1.16 | 0.91 | 0.77-1.08 |
| **Unmet need for contraception** |  |  |  |  |  |  |  |  |
| No | 1 |  | 1 |  | 1 |  | 1 |  |
| Yes | 3.94**<br>* | 3.45-4.50 | 2.12*** | 1.86-2.43 | 3.24*** | 2.74-3.85 | 2.28*** | 1.89-2.76 |

Ref: Reference group; Statistical significance: \* $p < 0.05$ , \*\* $p < 0.01$ , \*\*\* $p < 0.001$ .

Mothers who had secondary level education, were more likely to have unintended pregnancy than mothers who did not have any formal education in 2007 (OR: 1.28, 95% CI: 1.01-1.63), whereas in 2011, mothers with secondary level education were less likely to have unintended pregnancy than those who had no formal education (OR: 0.75, 95% CI: 0.60-0.95).

Employed mothers were more likely to have unintended pregnancy in 2007 (OR: 1.19, 95% CI: 1.04-1.37), 2011 (OR: 1.31, 95% CI: 1.12-1.53) and 2014 (OR: 1.19, 95% CI: 1.02-1.39) and Muslim mothers were more likely to have unintended pregnancy in 2011 (OR: 1.53, 95% CI: 1.26-1.86) and 2014 (OR: 1.33, 95% CI: 1.02-1.73). However, in 2017-18, Muslim mothers were less likely to have unintended pregnancy than mothers with Hindu and others religious faiths (OR: 0.73, 95% CI: 0.56-0.95).

Findings suggest that mothers from middle, richer and richest wealth quintiles in 2011 were less likely to have unintended pregnancy than those from the poorest quintile (OR: 0.81, 95% CI: 0.67-0.96 and OR: 0.82, 95% CI: 0.69-0.97, OR: 0.64, 95% CI: 0.52-0.79, respectively) and mothers from richest wealth quintile in 2017-18 were less likely to have unintended pregnancy than those from poorest (OR: 0.62, 95% CI: 0.47-0.82).

Mothers from households having a woman head were less likely to have unintended pregnancy in 2014 (OR: 0.74, 95% CI: 0.58-0.94) and 2017-18 (OR: 0.70, 95% CI: 0.57-0.87). Mothers whose families were comprising more than four members, were more likely to have unintended pregnancy in 2011, 2014 and 2017-18 BDHS.

In all four surveys, mothers whose age at first birth was more than 18 years were less likely to have unintended pregnancy than those whose age at birth of first child was less than 18 years (OR: 0.72, 95% CI: 0.63-0.81 in 2007; OR: 0.70, 95% CI: 0.63-0.78 in 2011; OR: 0.69, 95% CI: 0.60-0.79 in 2014 and OR: 0.73, 95% CI: 0.64-0.85 in 2017-18). In 2007, 2011 and 2017-18, the rural mothers were less likely to have unintended pregnancy than urban mothers (OR: 0.69, 95% CI: 0.58-0.81; OR: 0.77, 95% CI: 0.67-0.89 and OR: 0.72, 95% CI: 0.62-0.85, respectively in 2007, 2011 and 2017-18).

In 2007 and 2011, mothers from Sylhet division were less likely (OR: 0.68, 95% CI: 0.52-0.90 and OR: 0.66, 95% CI: 0.52-0.83, respectively) to have unintended pregnancy compared to those from Barisal division. Besides, in 2011 and 2017-18, mothers from Chittagong division were less likely (OR: 0.75, 95% CI: 0.60-0.93, and OR: 0.62, 95% CI: 0.47-0.82, respectively) to have unintended pregnancy compared to those from Barisal division. However, in 2014, mothers from Khulna and Rajshahi division were more likely (OR: 1.33, 95% CI: 1.01-1.74 and OR: 1.37, 95% CI: 1.04-1.79, respectively) to have unintended pregnancy than mothers from Barisal division.

Mothers who had unmet need for contraception during all four surveys were more likely to experience unintended pregnancy than those having their needs for contraception fulfilled (OR: 3.94, 95% CI: 3.45-4.50 in 2007; OR: 2.12, 95% CI: 1.86-2.43 in 2011; OR: 3.24, 95% CI: 2.74-3.85 in 2014 and OR: 2.28, 95% CI: 1.89-2.76 in 2017-18).

## Discussion

This large-scale comparative study focuses over measuring the prevalence of unintended pregnancies among ever married women aging 15-49 at different time points and exploring potential factors influencing the trend of such pregnancies in Bangladesh from 2007 to 2018 as per last four BDHSs. Our results show that the prevalence of unintended pregnancy has attained a substantial decrease from 29% to about 21% over the last decade, compared to the spell between 1993 and 2007 when the prevalence decreased from 33% to 29%. Despite significant improvement, it’s still leaving room for progress through more curtailed interventions.

A wide gamut of socio-demographic predictors, which were chosen based on literature reviews of multiple similar studies across developing countries, has been investigated in our study to identify their impacts on occurrence of unintended pregnancies among ever married women. The factors found to have statistically significant associations with the tendency of any unintended pregnancy were age, employment status, wealth quintile, education, religion, division, family size, gender of household head, age at first birth, birth order and unmet need for contraception. And many of these factors draw need for programmatic implementations.

Our findings reported that women who were above 18 years of age at the time of their first birth presented lower odds of unintended pregnancies compared to their counterparts. Usually, mothers aged more than18 years are likely to be more aware about sexual and reproductive health whereas under 18 mothers have inadequate understanding in this domain and are more likely to experience unintended pregnancies within the 6 months of first childbirth [28]. Also, mothers facing unmet need of contraception were more likely to go through unintended pregnancies than the mothers whose need for contraception were fulfilled. It’s a possibility that women getting married at earlier ages have less knowledge about contraception and hence are exposed to risks of experiencing unintended pregnancies. Another reason could be that adolescent mothers have a very limited access to practicing contraception in early marital life [29, 30]. Findings from similar studies conducted in Nepal, Japan, Angola and one recent study from Bangladesh portray the same exact findings [19, 29, 31, 32], which are testaments for an extreme necessity of strict prevention of underage marriages as per government legislatures and availing mothers with more frequent opportunities of exercising contraception particularly in early marital life.

We found that, women’s increasing age had a negative association with unintended pregnancies and young adolescents were more susceptible to conceive unplanned. One possible explanation to the findings might be young women’s desire to have some years of interval for first pregnancy preparation, but due to lack of exposure to favourable contraceptives and cultural influences, they end up having mistimed conceptions [29, 30]. However, the findings are in total contrast with several other studies conducted in Nigeria, Iran, Nepal and Vietnam where women with advancing age particularly those being 35 years and above displayed higher odds of unintended pregnancies [29, 33-35]. Also, women belonging to higher wealth quintiles were less susceptible to unplanned conceptions than those being in poorer economic conditions. This finding aligns with multiple studies portraying evidences of association between socio-economic status and unintended pregnancies as women with limited financial abilities can often face the impetus of unwarranted events in their life [20, 35, 36]. Hence, particular emphasis should be given upon poor and younger women in programmatic implementations to reduce this burden.

Women from female headed households were found to be less prone to unintended pregnancies. Studies also suggest that decision making opportunities in the household provide women an enhanced sense of autonomy and thus helps taking intellectual steps around the key events of life like that of birth control [37, 38]. However, employed and urban women had higher odds of having unintended pregnancies and the trend is similar over the period of time. Though the relationship is opposite to another two census based studies conducted in USA and Kenya where employed and urban women were less likely to have unintended conceptions [39, 40]. This might have happened due to dissimilarity in socio-economic contexts between the two countries. More rigorous studies are required to further examine this variable.

Our study revealed that women having higher number of parities (birth order 1+) and consequentially with larger family sizes displayed more likeliness of having unintended pregnancies. Other regional studies conducted at Kenya, Ethiopia and China also had similar findings [40-43]. The patriarchal traits of desiring male child over female particularly embedded in South Asian rural cultures have been proved to increase parity progression in previous studies [44-46], which might lead to unintended pregnancies. Also, contextually, the desires for building a larger family might also contribute to unintended pregnancies [33]. However, BDHS doesn’t collect data on the cultural factors influencing reproductive behaviour of the families. However, these findings cast light upon the utmost need of ensuring access to fundamental, proper and close-to-door maternal care services for every mother in the region.

The main strength of this study has been the use of nationally representative data from recent four consecutive demographic health surveys. Whereas, there are certain limitations, such as; the information collected through the cross-sectional design of the surveys doesn’t guarantee any potential predictors of unintended pregnancy. Secondly, unintended pregnancies were determined based on self-reported data and can be subjected to recall bias and degree of perception of the participant mothers. Besides, some crucial variables might have been overlooked while testing for potential impact on unintended pregnancy due to absence of adequate literatures in favour.

## Conclusion

The prevalence of unintended pregnancy declined in the last decade though it still remains a serious problem for women in Bangladesh and is declining at a slow rate. We highlighted several factors attaining heightened influence on unintended pregnancy. Policy makers and healthcare providers can benefit from the evidences generated in this study and can cater their services to provide support for women from poorer section, women with larger family sizes and underaged adolescent girls and empower them to avail regular reproductive and maternal healthcare services with particular focus on contraception preferences, to curb the incidence of unintended pregnancies. Also, further studies are required to minutely assess the impact of urban culture and employment occupied lifestyle on women’s decision making around pregnancy, to ensure the wellbeing of mothers and children from all walks of the society.

## Data Availability

Data used for analysis in this paper are available in Bangladesh Demographic and Health Surveys 2007, 2011, 2014 and 2018

## Abbreviations

BDHS: Bangladesh demographic and Health Survey
CI: Confidence Interval
EA: Enumeration Area
icddr,b: International Centre for Diarrheal Disease Research, Bangladesh
NIPORT: National Institute of Population Research and Training
OR: Odds Ratio
PSU: Primary Sampling Unit
SDG: Sustainable Development Goal
TWFR: Total Wanted Fertility Rate

## Acknowledgments

The authors want to sincerely thank all females participating in 2007, 2011, 2014 and 2017-18 Bangladesh Demographic and Health Surveys.

## References

[1] G. Barrett and K. Wellings, “What is a ‘planned’pregnancy? Empirical data from a British study,” Social science & medicine, vol. 55, no. 4, pp. 545–557, 2002.

[2] World Health Organization, “Unwanted Pregnancies and the Need to Increase Access to Modern Contraceptives: Factsheet,” Reproductive, Maternal, Newborn, Child and Adolescent Health Division of NCD and Health through the Life-Course, World Health Organization, Regional Office for the Western Pacific, 2018. [Online]. Available: http://www.wpro.who.int/reproductive_health/factsheets/en/

[3] J. Bearak, A. Popinchalk, L. Alkema, and G. Sedgh, “Global, regional, and subregional trends in unintended pregnancy and its outcomes from 1990 to 2014: estimates from a Bayesian hierarchical model,” The Lancet Global Health, vol. 6, no. 4, pp. e380-e389, 2018.

[4] United Nations, “Trends in Contraceptive Use Worldwide 2015,” Department of Economic and Social Affairs, Population Division Newyork, 2015. [Online]. Available: http://www.un.org/en/development/desa/population/publications/pdf/family/trendsContraceptiveUse2015Report.pdf

[5] E. Zuehlke, “Reducing unintended pregnancy and unsafely performed abortion through contraceptive use,” 2009.

[6] S. Abbasi, C. H. Chuang, R. Dagher, J. Zhu, and K. Kjerulff, “Unintended pregnancy and postpartum depression among first-time mothers,” Journal of Women’s Health, vol. 22, no. 5, pp. 412–416, 2013.

[7] P. S. Shah, T. Balkhair, A. Ohlsson, J. Beyene, F. Scott, and C. Frick, “Intention to become pregnant and low birth weight and preterm birth: a systematic review,” Maternal and child health journal, vol. 15, no. 2, pp. 205–216, 2011.

[8] J. S. Taylor and H. J. Cabral, “Are women with an unintended pregnancy less likely to breastfeed?,” Journal of family practice, vol. 51, no. 5, pp. 431–437, 2002.

[9] P. J. Surkan et al., “Unintended pregnancy is a risk factor for depressive symptoms among socio-economically disadvantaged women in rural Bangladesh,” BMC pregnancy and childbirth, vol. 18, no. 1, pp. 1–13, 2018.

[10] M. A. Habib et al., “Prevalence and determinants of unintended pregnancies amongst women attending antenatal clinics in Pakistan,” BMC pregnancy and childbirth, vol. 17, no. 1, pp. 1–10, 2017.

[11] National Institute of Population Research and Training (NIPORT). and ICF International., “Bangladesh Demographic and Health Survey 2017-18,” Dhaka, Bangladesh, and Rockville, Maryland, USA: NIPORT, and ICF, 2020.

[12] K. Santhya, “Early marriage and sexual and reproductive health vulnerabilities of young women: a synthesis of recent evidence from developing countries,” Current opinion in obstetrics and gynecology, vol. 23, no. 5, pp. 334–339, 2011.

[13] World Health Organization, “Global and regional estimates of the incidence of unsafe abortion and associated mortality in 2008,” Geneva: WHO, 2011.

[14] T. K. Roy, M. N. Huda, B. P. Singh, and K. Singh, “Prevalence of socioeconomic correlates of unplanned pregnancy in Bangladesh,” International Journal of Research in Economics and Social Sciences, vol. 5, no. 6, pp. 183–195, 2015.

[15] K. R. Tapan, D. Sarkar, and B. Singh, “Risk factors of unplanned pregnancy for reproductive health in Bangladesh: A case study”,” Journal of Science and Technology, vol. 4, no. 1, pp. 1–10, 2014.

[16] A. O. Tsui, R. McDonald-Mosley, and A. E. Burke, “Family planning and the burden of unintended pregnancies,” Epidemiologic reviews, vol. 32, no. 1, pp. 152–174, 2010.

[17] G. Bishwajit, S. Tang, S. Yaya, and Z. Feng, “Unmet need for contraception and its association with unintended pregnancy in Bangladesh,” BMC pregnancy and childbirth, vol. 17, no. 1, p. 186, 2017.

[18] S. M. I. Hossain, M. Khan, M. Rahman, and M. P. Sebastian, “South east Asia regional training manual,” New Delhi, India: Population Council, 2005.

[19] G. Bishwajit, S. Tang, S. Yaya, and Z. Feng, “Unmet need for contraception and its association with unintended pregnancy in Bangladesh,” BMC pregnancy and childbirth, vol. 17, no. 1, pp. 1–9, 2017.

[20] M. Kamal and A. Islam, “Prevalence and socioeconomic correlates of unintented pregnancy among women in rural Bangladesh,” salud pública de méxico, vol. 53, no. 2, pp. 108–115, 2011.

[21] F. A. Huda, A. Ahmed, H. R. Mahmood, F. Ahmmed, A. Panza, and R. Somrongthong, “Delaying first pregnancy in reducing burden of unintended pregnancy among married adolescents in urban slums of Bangladesh: A situation analysis,” Journal of Health Research, 2018.

[22] F. R. Noor, M. M. Rahman, U. Rob, and B. Bellows, “Unintended pregnancy among rural women in Bangladesh,” International quarterly of community health education, vol. 32, no. 2, pp. 101–113, 2012.

[23] Alfredo Aliaga & Ruilin Ren, “Optimal Sample Sizes for Two-stage Cluster Sampling in Demographic and Health Surveys, No. 30.” [Online]. Available: https://dhsprogram.com/pubs/pdf/WP30/WP30.pdf

[24] M. a. A. National Institute of Population Research and Training (NIPORT), and Macro International.,, “Bangladesh Demographic and Health Survey, 2007,” Dhaka, Bangladesh and Calverton, Maryland, USA, 2009.

[25] National Institute of Population Research and Training (NIPORT)., Mitra and Associates., and ICF International., “Bangladesh Demographic and Health Survey 2011,” NIPORT, Mitra and Associates, and ICF International, Dhaka, Bangladesh and Calverton, Maryland, USA, 2012.

[26] National Institute of Population Research and Training (NIPORT)., Mitra and Associates., and ICF International., “Bangladesh Demographic and Health Survey 2014,” NIPORT, Mitra and Associates, and ICF International, Dhaka, Bangladesh and Calverton, Maryland, USA, 2015.

[27] D. W. Hosmer Jr, S. Lemeshow, and R. X. Sturdivant, Applied logistic regression. John Wiley & Sons, 2013.

[28] A. J. Knutson, S. S. Boyd, J. B. Long, and K. H. Kjerulff, “Early resumption of sexual intercourse after first childbirth and unintended pregnancy within six months,” Women’s Health Issues, vol. 32, no. 1, pp. 51–56, 2022.

[29] R. Adhikari, K. Soonthorndhada, and P. Prasartkul, “Correlates of unintended pregnancy among currently pregnant married women in Nepal,” BMC International Health and Human Rights, vol. 9, no. 1, pp. 1–10, 2009.

[30] S. A. A. Aziz Ali, S. Aziz Ali, and N. S. Khuwaja, “Determinants of unintended pregnancy among women of reproductive age in developing countries: a narrative review,” Journal of Midwifery and Reproductive Health, vol. 4, no. 1, pp. 513–521, 2016.

[31] A. Goto, S. Yasumura, M. R. Reich, and A. Fukao, “Factors associated with unintended pregnancy in Yamagata, Japan,” Social science & medicine, vol. 54, no. 7, pp. 1065-1079, 2002.

[32] S. Yaya and B. Ghose, “Prevalence of unmet need for contraception and its association with unwanted pregnancy among married women in Angola,” PloS one, vol. 13, no. 12, p. e0209801, 2018.

[33] L. Cu Le, R. Magnani, J. Rice, I. Speizer, and W. Bertrand, “Reassessing the level of unintended pregnancy and its correlates in Vietnam,” Studies in family planning, vol. 35, no. 1, pp. 15–26, 2004.

[34] M. J. Abbasi-Shavazi, M. Hosseini-Chavoshi, A. Aghajanian, B. Delavar, and A. Mehryar, “Unintended pregnancies in the Islamic Republic of Iran: levels and correlates,” Asia-Pacific population journal, vol. 19, no. 1, pp. 27–38, 2004.

[35] G. Sedgh, A. Bankole, B. Oye-Adeniran, I. F. Adewole, S. Singh, and R. Hussain, “Unwanted pregnancy and associated factors among Nigerian women,” International family planning perspectives, pp. 175–184, 2006.

[36] N. Jaeni, P. McDonald, and I. Utomo, “Determinants of unintended pregnancy among ever-married women in Indonesia: An analysis of the 2007 IDHS,” Australian Demographic and Social Research Institute, 2009.

[37] R. Yogendrarajah, “Women empowerment through decision making,” *Yogendrarajah*, Rathiranee, (2013), Women Empowerment through Decision Making, The International Journal of Economics and Business Management, vol. 3, no. 1, 2013.

[38] M. Rahman, “Women’s autonomy and unintended pregnancy among currently pregnant women in Bangladesh,” Maternal and child health journal, vol. 16, no. 6, pp. 1206-1214, 2012.

[39] J. H. Su, “Local employment conditions and unintended pregnancy,” Journal of Marriage and Family, vol. 81, no. 2, pp. 380–396, 2019.

[40] L. Ikamari, C. Izugbara, and R. Ochako, “Prevalence and determinants of unintended pregnancy among women in Nairobi, Kenya,” BMC pregnancy and childbirth, vol. 13, no. 1, pp. 1–9, 2013.

[41] E. A. Kassahun et al., “Factors associated with unintended pregnancy among women attending antenatal care in Maichew Town, Northern Ethiopia, 2017,” BMC research notes, vol. 12, no. 1, pp. 1-6, 2019.

[42] H. Wang, et al., “Contraception and unintended pregnancy among unmarried female university students: a cross-sectional study from China,” PloS one, vol. 10, no. 6, p. e0130212, 2015.

[43] B. Hamdela, A. G/mariam, and T. Tilahun, “Unwanted pregnancy and associated factors among pregnant married women in Hosanna Town, Southern Ethiopia,” PloS one, vol. 7, no. 6, p. e39074, 2012.

[44] S. Chaudhuri, “The desire for sons and excess fertility: a household-level analysis of parity progression in India,” International perspectives on sexual and reproductive health, pp. 178–186, 2012.

[45] R. Bairagi, “Effects of sex preference on contraceptive use, abortion and fertility in Matlab, Bangladesh,” International Family Planning Perspectives, pp. 137–143, 2001.

[46] M. Das Gupta, J. Zhenghua, L. Bohua, X. Zhenming, W. Chung, and B. Hwa-Ok, “Why is son preference so persistent in East and South Asia? A cross-country study of China, India and the Republic of Korea,” The Journal of Development Studies, vol. 40, no. 2, pp. 153–187, 2003.

